# A poor-man’s approach to the effective reproduction number: the COVID-19 case

**DOI:** 10.1101/2020.04.22.20076430

**Authors:** José Menéndez

## Abstract

It is shown that estimates of the effective reproduction number *R*_*t*_ for COVID-19 using standard packages such as EpiEstim can be reproduced very accurately using the expression *R*_*t*_ = *c*(*t*)/*c*(*t* − *τ*), where *c*(*t*) is the incidence at time *t* and *τ* the mean value of the series interval.

---

The effective reproduction number *R*_*t*_ —a fundamental epidemiological parameter that characterizes the temporal dynamics of an infectious disease—is notoriously difficult to determine without detailed modeling[1-6]. During the ongoing COVID-19 pandemic, daily tallies of the incidence *c*(*t*) (new cases on day *t*), recovered *r*(*t*) (individuals who are declared cured on day *t*) and deceased *d*(*t*) (individuals who die on day *t*) have become available for many countries and regions [7]. With this information, one might naively expect that *R*_*t*_ can be determined as

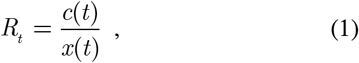

with *x*(*t*) = *r*(*t*) + *d*(*t*). This definition is exactly equivalent to the expression for the basic reproduction number in a ‘Susceptible-Infectious-Removed’ (SIR) framework, but can be obviously applied to the observed data without any reference to a specific model. Calculations with Eq. (1) for COVID-19, however, disagree with values of *R* computed from standard packages such as EpiEstim [4], as seen in Fig. 1.

**FIG. 1.**
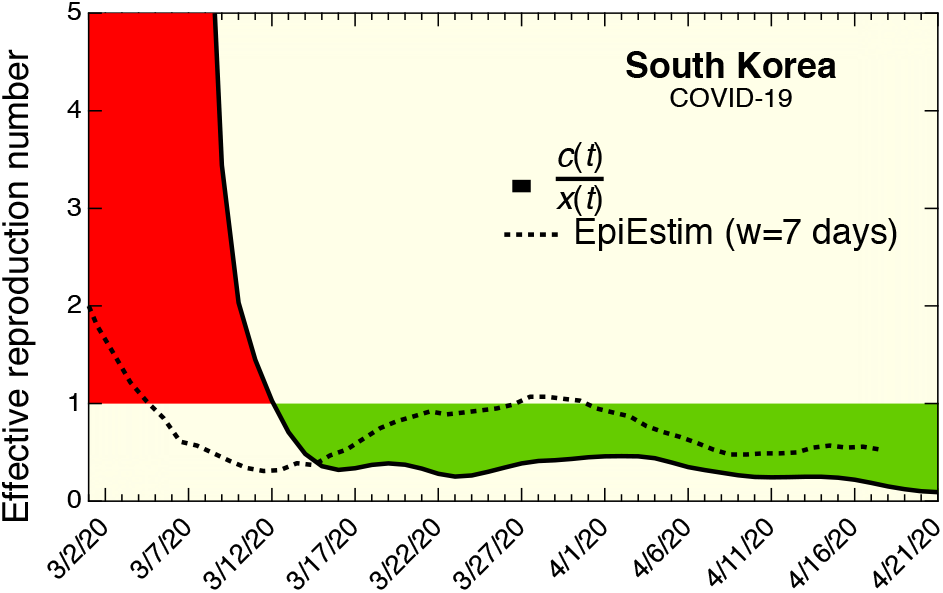
Solid black line: effective COVID-19 reproduction number *R*_*t*_ for South Korea calculated from Eq. (1). Dotted line: *R*_*t*_ calculated with the package EPIESTIM using a mean infectious Period *τ* = 5.8 days with a standard deviation of 2.9 days and a time window of *w* = 7 days.

Part of the reason for the discrepancy is the lack of reliable *r*(*t*) data. For some countries, such as the United Kingdom, the published cumulative number of recovered people is *less* than the cumulative number of deceased individuals, a very unlikely scenario. Even globally, the sum total of deceased individuals is a substantial fraction of cured individuals, which is also highly implausible. Furthermore, there is a *relative* time shift between *c*(*t*) and *x*(*t*), since the former includes the incubation time plus the time to develop symptoms serious enough to be reported. However, attempting to correct for this relative delay does not improve the agreement in Fig. 1 in any significant way.

Standard packages do not suffer from the above problems because they rely only on *c*(*t*) data, and therefore are not affected by systematic errors in *r*(*t*). For these calculations one requires the infectivity profile of the disease, which is approximated as the distribution of the standard interval [4]. Calculations based on EpiEstim [4] using a series interval mean *τ* = 4.7 days and standard deviation 2.9 days, from Ref. [8], are updated daily at [9]. A more recent study finds *τ* = 5.8 days, with 44% presymptomatic transmission [10]. The recovery time, on the other hand, is much longer, and this implies that individuals at advanced stages of the disease have a very low infectivity. Under these conditions Eq. (1) cannot be valid, since its denominator should not be *x*(*t*) but the number of people who became infected approximately at time *t* − *τ*. The latter is greater than the former during the ramp-up phase of the disease, and this explains why the curve calculated from Eq. (1) shows a higher *R*_*t*_ during this time. Conversely, the number of people who became infected at time *t* − *τ* is *less* than *x*(*t*) at later stages, and this explains why the curve calculated from Eq. (1) gives a lower value of the number of *R*_*t*_ at these times.

The above considerations suggest an alternative definition of *R*_*t*_ as

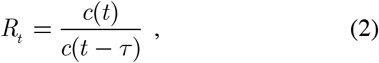

A computation of *R*_*t*_ for different countries using Eq. (2) with *τ* = 5.8 days is shown in Figs. 2-5, where it is compared with calculations of *R*_*t*_ using EpiEstim for the same value of *τ*. Since the *c*(*t*) data are quite noisy, a smooth version *c*_*s*_ (*t*) is used. The smooth *c*_*s*_ (*t*) is computed using a regularization approach that minimizes the sum of the mean square deviation between *c*_*s*_ (*t*) and *c*(*t*) plus a term proportional to the square of the second derivative *c*_*s*_′′(*t*). The relative weight of the two terms is controlled by a parameter *λ* that is defined as in Eq. 3 of Ref. [11], with the matrices **B** and 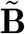 taken as the identity matrix. The chosen value was *λ* =100 day^4^. A comparison between *c*_*s*_ (*t*) and *c*(*t*) is shown in the inset of Fig. 2 for the case of South Korea. The regularization method has the advantage that it distorts the shape of a curve less than a typical sliding average, while being superior to Savitzky-Golay[12] smoothing in terms of noise reduction. This was important in the context of the comparison made here between Eq. (2) and the EpiEstim package, because artificial shape distortions are eliminated. For practical applications of Eq. (2), however, a simple 7- day running average should suffice.

**FIG. 2.**
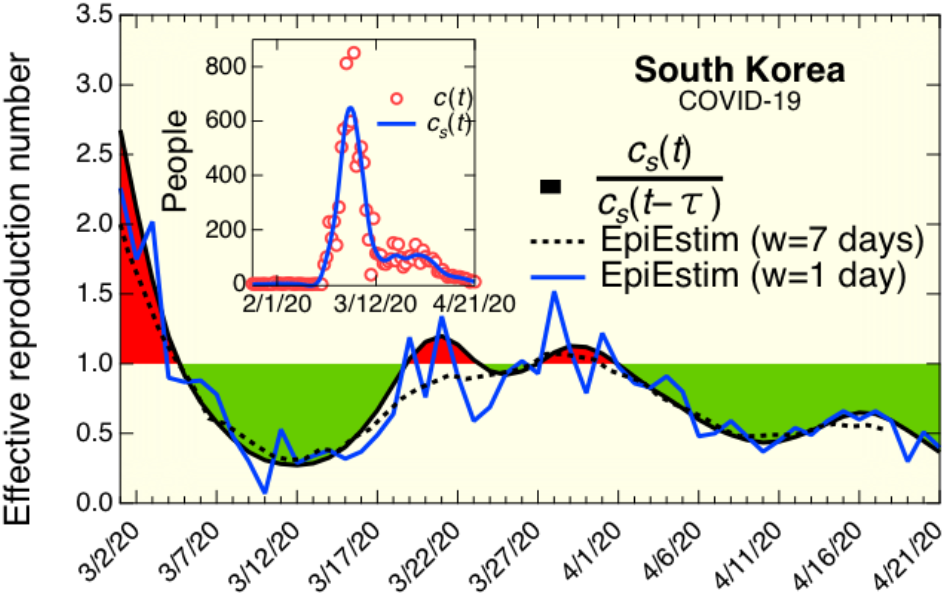
The COVID-19 effective reproduction number for South Korea. The solid black line is from Eq. (2). The blue and dotted black lines are estimates from EPIESTIM using time windows of 1 day and 7 days, respectively. All calculations used *τ* = 5.8 days, with a standard deviation (EPIESTIM case) of 2.9 days. The inset shows the reported incidence *c*(*t*) (red circles) and the smoothed curve *c*_*s*_ (*t*).

**FIG. 3.**
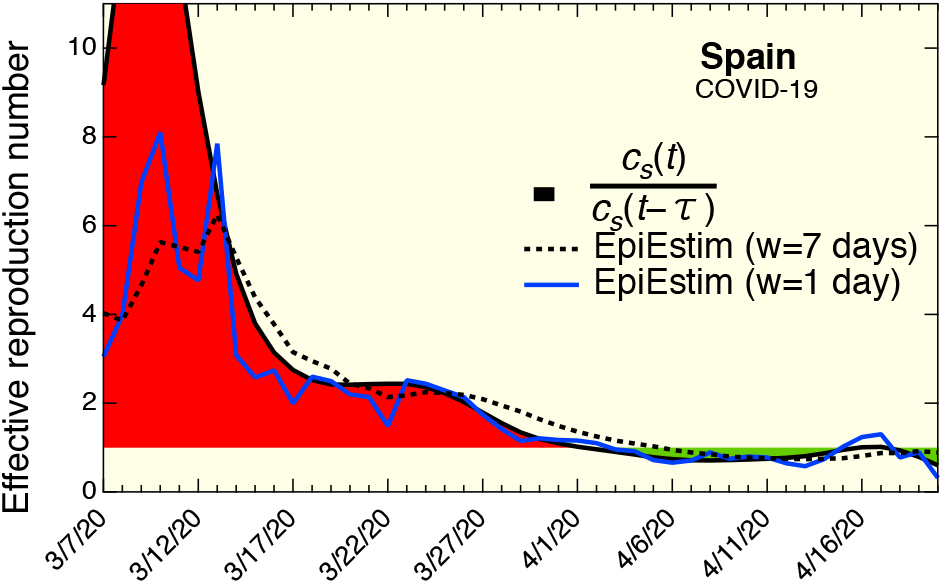
The COVID-19 effective reproduction number for Spain. The solid black line is from Eq. (2). The blue and dotted black lines are estimates from EPIESTIM using time windows of 1 day and 7 days, respectively. All calculations used *τ* = 5.8 days, with a standard deviation (EPIESTIM case) of 2.9 days.

**FIG. 4.**
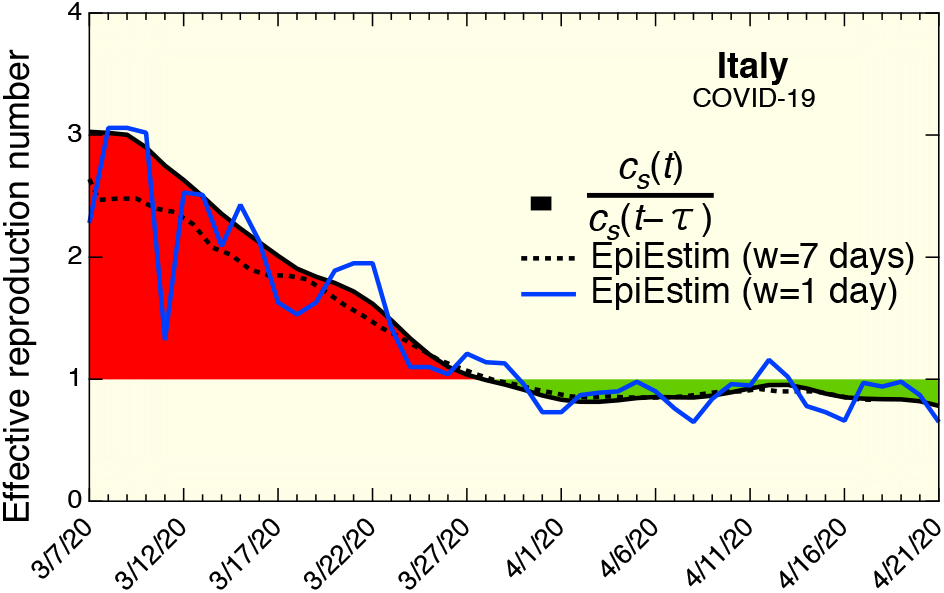
The COVID-19 effective reproduction number for Italy. The solid black line is from Eq. (2). The blue and dotted black lines are estimates from EPIESTIM using time windows of 1 day and 7 days, respectively. All calculations used *τ* = 5.8 days, with a standard deviation (EPIESTIM case) of 2.9 days.

**FIG. 5.**
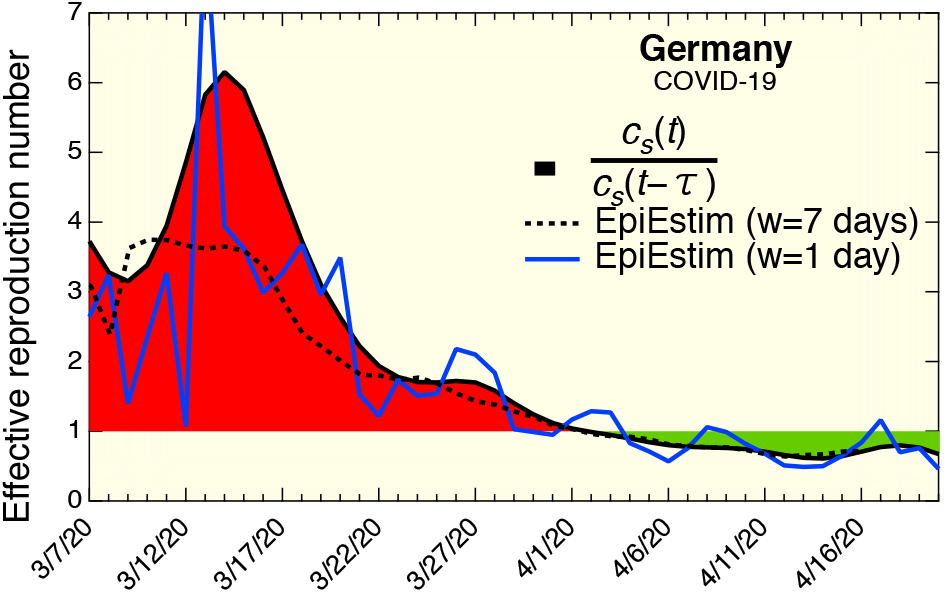
The COVID-19 effective reproduction number for Germany. The solid black line is from Eq. (2). The blue and dotted black lines are estimates from EPIESTIM using time windows of 1 day and 7 days, respectively. All calculations used *τ* = 5.8 days, with a standard deviation (EPIESTIM case) of 2.9 days.

**FIG. 6.**
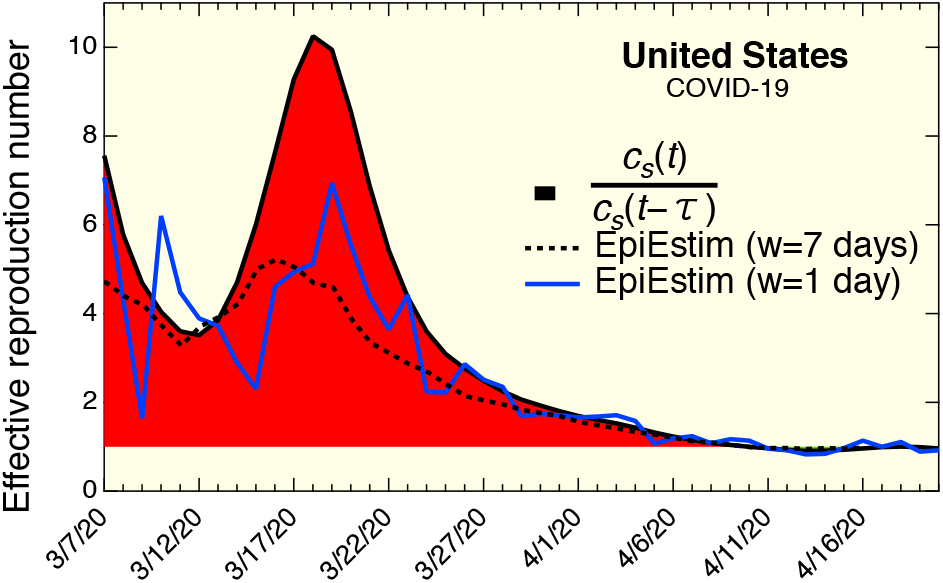
The COVID-19 effective reproduction number for the United States. The solid black line is from Eq. (2). The blue and dotted black lines are estimates from EPIESTIM using time windows of 1 day and 7 days, respectively. All calculations used *τ* = 5.8 days, with a standard deviation (EPIESTIM case) of 2.9 days.

**FIG. 7.**
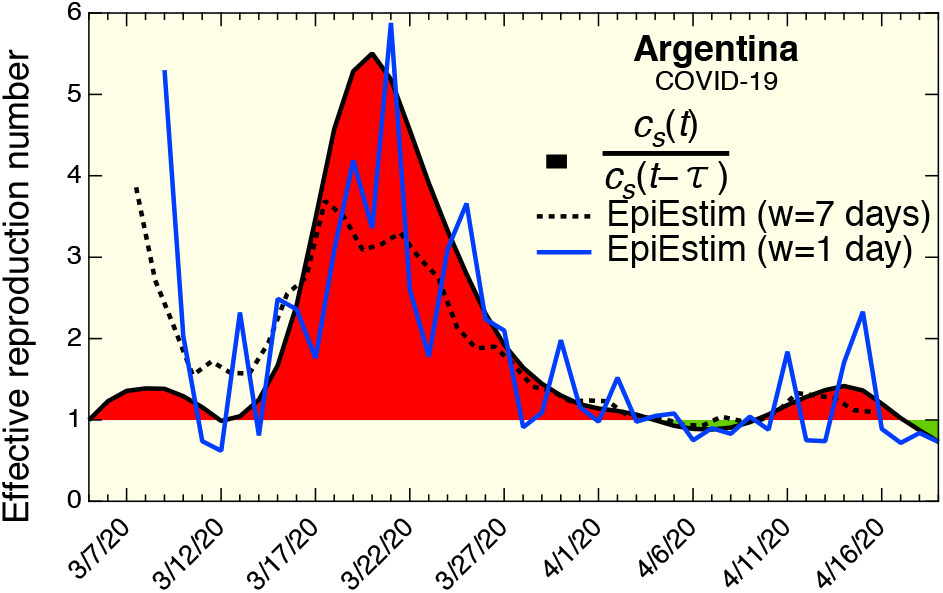
The COVID-19 effective reproduction number for Argentina. The solid black line is from Eq. (2). The blue and dotted black lines are estimates from EPIESTIM using time windows of 1 day and 7 days, respectively. All calculations used *τ* = 5.8 days, with a standard deviation (EPIESTIM case) of 2.9 days.

Two calculations with EpiEstim were performed, both with *τ* = 5.8 days and a standard deviation of 2.9 days. The first calculation uses a time window *w* = 1 day, and the second calculation a time window *w* =7 days. In the latter case, the value that appears in the figures at a particular time *t* corresponds to the window for which *t* is the middle point. The *w* = 1 day results are very noisy, as expected, but both EpiEstim curves are in very good agreement with the calculations from Eq. (2). It is important to point out that the incidence data have a significant delay with respect to the actual time of infection, so that all figures should be interpreted at representing the situation between one and two weeks before the dates in the horizontal axes. We made no attempt to correct for reporting delays as in Ref. [9].

In summary, an extremely simple algorithm has been introduced that yields effective reproduction numbers very similar to those obtained from more elaborate approaches as exemplified by the EpiEstim package. The latter should produce more accurate results if the full epidemiological characteristics of the disease are well known, but for calculations at the present time when little is known about COVID-19, the method proposed here is much easier to use and provides comparable accuracy

The numerical simulations shown here were carried out on Igor Pro 8.0 (Wavemetrics, Inc). The code is available upon request.

## Data Availability

Data and underlying code are available upon request.

## Notes

### Competing Interest Statement

The authors have declared no competing interest.

### Funding Statement

No external funding.

